# Towards Accessible Radiological Image Analysis via Local Agentic Framework: Validation in Mammography

**DOI:** 10.64898/2026.08.03.26359608

**Authors:** Lejun Cen, Jessica H. Porembka, Jody C. Hayes, Kanwal Merchant, Uzoma Igboagi, Shweta Srivastava, Ann R. Mootz, Viktoria Topper, Sterling Hayes, Neha Yadu, Firouzeh K. Arjmandi, Jennifer G. Schopp, Basak E. Dogan, Tianshen Hu

**Author notes:** These authors jointly supervised this work and are designated as co-last authors (Basak E. Dogan, Tianshen Hu).

## Abstract

Existing radiological artificial intelligence (AI) systems are difficult to modify, validate, and adapt to new clinical applications. We present a large language model (LLM)-driven agentic framework capable of reconstructing, optimizing, and customizing deep-learning (DL) systems for radiological image analysis using a single consumer-grade PC. The agent reconstructed the missing pre-training model and corrected a clinical reasoning flaw in an example mammography DL workflow. The improved model performance surpassed all 1,687 submitted models in the Radiological Society of North America Breast Cancer AI Challenge. Across international datasets (n>13,000) from US and China, the model demonstrated robust generalizability (AUC: 0.9). In a reader study (n>1,200), the model outperformed radiologists by an absolute AUC margin of 24% on extended follow-up. Our findings demonstrate that LLM-driven agents enable radiologist-guided customization of radiological AI systems on a consumer-grade PC while reducing the technical expertise required for implementation. This work paves the way for accessible radiological AI.

## 1. Introduction

Radiological screening programs are among the most effective strategies for reducing cancer mortality through early detection ^1,2^. For instance, breast cancer remains the second leading cause of cancer-related death among women ^3–5^, and screening mammography has been estimated to account for approximately 25% of breast cancer deaths averted ^1^. Accordingly, many countries have implemented large-scale breast cancer screening programs. In the US alone, over 40 million screening mammograms are performed each year at a cost of over $10 billion annually ^6,7^. However, the efficacy of these programs is limited by the inherent challenges of mammographic interpretation. Radiologists exhibit inter-reader variability and face limitations in sensitivity and specificity ^8–10^, leading to false positives that may cause patient anxiety and invasive procedures^11–13^, or false negatives that may allow cancers to progress to advanced stages ^14^.

Artificial intelligence (AI) may offer a promising avenue to improve accuracy ^15,16^ and mitigate the global shortage of subspecialty-trained radiologists ^17–19^. While early generations of computer-aided detection (CAD) failed to deliver real-world performance gains ^20–25^, the current “renaissance” in deep-learning (DL) has produced systems with standalone performance comparable to human readers ^13,26–29^. McKinney et al. (2020) demonstrated a DL system that claimed to surpass the performance of individual radiologists across large-scale datasets from both the UK and the US ^28^. More recently, Eisemann et al. (2025) reported that AI-supported double reading in Germany increased cancer detection by 17.6% while decreasing recall rates by 2.5% ^13^. Similarly, Kelly et al. (2026) showed in a multicenter UK study that AI achieved higher sensitivity than human readers in over 100,000 women ^29^.

Although DL models show strong potential, their clinical integration, especially in developing countries, remains relatively limited due to high infrastructure costs, as well as lack of explainability and controllability. Most current proprietary DL systems function as “black boxes,” lacking the transparency essential for building clinician trust. Furthermore, radiologists are typically restricted to minor parameter adjustments, leaving them unable to modify core model logic or tailor systems to specific clinical settings and diverse patient demographics. Customizing or fundamentally modifying these DL tools currently requires a multi-disciplinary collaboration among radiologists, AI experts, and programmers, which is a resource unavailable to most radiologists.

The maturation of Large Language Models (LLMs), in particular “reasoning LLMs” in 2024 and 2025, and autonomous AI agents is redefining human-AI collaboration and yielding significant leaps in programming and problem-solving abilities ^30^, including for medical tasks ^31^. In radiology, LLMs have successfully passed radiology board–style examinations and achieved 88% accuracy on radiology examination questions for medical trainees, compared to 76% accuracy for medical students ^32,33^. A key milestone is DeepSeek R1, the first open-source reasoning model with capabilities comparable to proprietary alternatives. Unlike standard LLMs, these models perform multi-step reasoning and can provide step-by-step explanations of their outputs ^34,35^.

LLM-based agents are semi-autonomous systems that operate as interconnected multi-agent systems that can use various tools to manage complex workflows. In software engineering, coding agents have become increasingly sophisticated, capable of searching open-source repositories to generate customized software solutions ^36^. In the medical field, agents have been reported to achieve 87.5% accuracy in autonomous tool usage, whereby they are able to independently select and execute specialized tools such as histopathology analysis algorithms, medical image segmentation software, and queries to clinical databases ^37^.

Integrating agentic coding with medical reasoning can improve model explainability and controllability. However, existing radiological AI agents are largely confined to basic tasks that use tools directly without modifying their core logic, such as querying black-box proprietary DL tools for image analysis. Furthermore, many medical agents remain heavily reliant on cloud-based proprietary LLMs. Beyond high token costs, uploading sensitive patient data to proprietary cloud servers poses severe data privacy and security risks.

In this study, we propose an agentic framework for radiological image analysis built around three guiding principles: explainability, controllability, and accessibility (Figure 1A). The framework is grounded in an open-source ecosystem that provides complete transparency across all system components (LLM, agent, and DL tools). With full code transparency and LLM-generated explanations, radiologists can understand and therefore control the underlying AI logic to integrate their clinical expertise through agentic coding. Our proposed framework prioritizes accessibility by executing the entire AI system locally on a single consumer-grade PC, avoiding costly industrial workstations or cloud-related token fees and risks. Using screening mammography as a proof-of-concept, we demonstrated the viability of this accessible and controllable radiological AI workflow. We evaluated system performance across varying levels of agentic autonomy by repairing, optimizing, and customizing an open-source DL tool (Figures 1B,C and 2). Its generalizability was validated through a retrospective study on international mammography datasets alongside an independent reader study.

**Figure 1.**
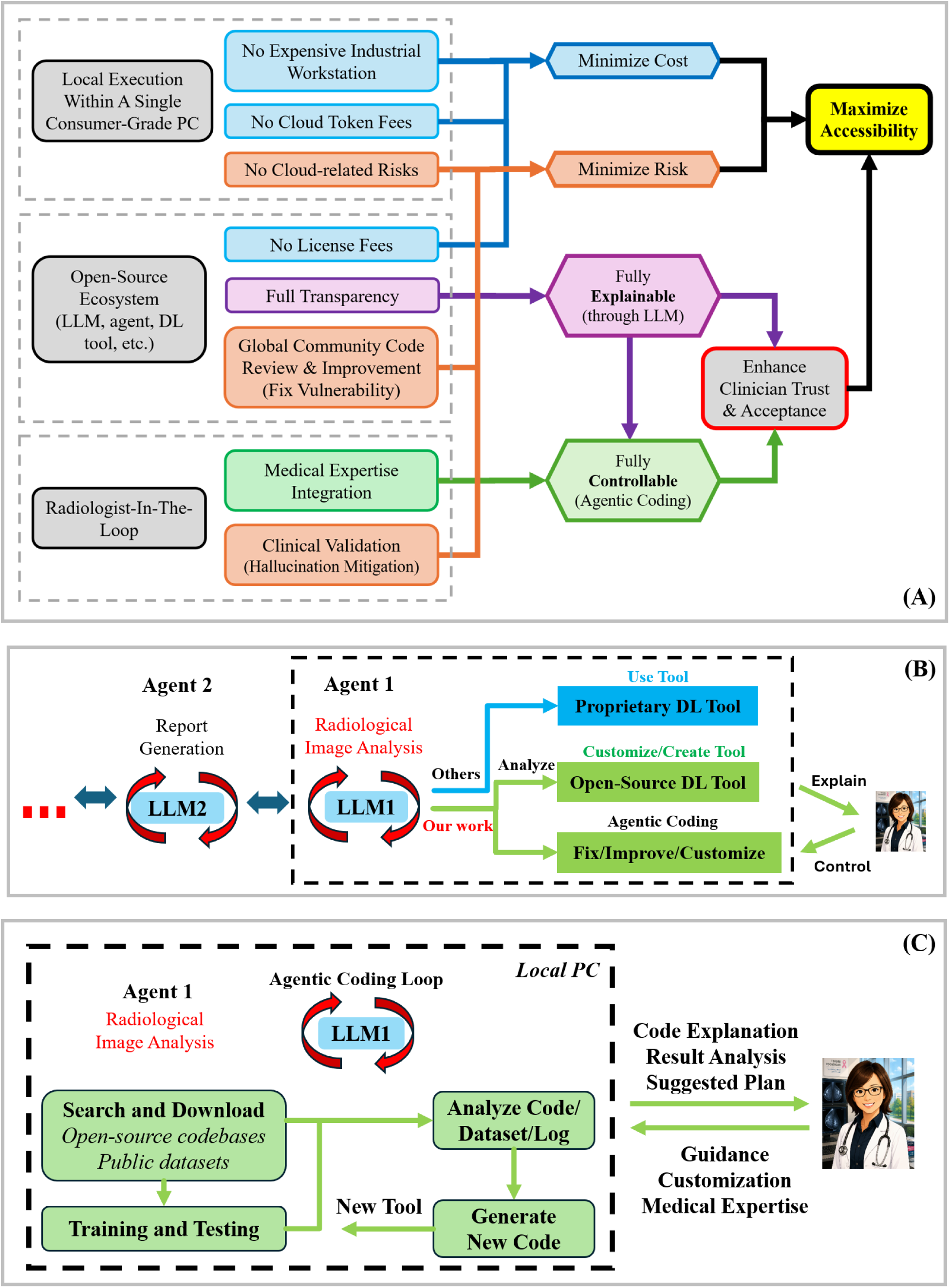
(A) Overview of the proposed LLM-driven agentic radiology framework. (B) In an example workflow, the system may consist of multiple agents, such as a radiological analysis agent (LLM1), a report-generation agent (LLM2), and others. Our study centers on the image-analysis component (in red). Conventional radiology agents typically rely on proprietary DL tools that operate as black boxes. Our LLM-driven framework addresses this limitation by using open-source DLs that can provide transparency for the LLM to analyze and explain the details at both the workflow level and model level to radiologists. This can help radiologists to use agentic coding to control the DL tools on different levels, such as fixing, improving, or customizing the core logic. (C) Autonomous agent centered on an LLM that operates within an iterative loop (red circular arrows), continuously planning, evaluating, and refining its actions. The agent searches for and downloads external open-source codebases and public datasets. These inputs are incorporated into a training and testing stage, where their performance is evaluated. The agent can also conduct analysis of code, datasets, or logs. This information is delivered for expert evaluation (bidirectional arrows), allowing radiologists and/or other domain experts to provide guidance, validate intermediate or final results, and customize system behavior. The feedback is reintegrated into the workflow, forming a feedback cycle.

## 2. System Architecture and Design Principles

### 2.1 Explainability

Our proposed framework leverages the full transparency of open-source ecosystems and the multi-disciplinary knowledge of the reasoning LLM to provide interpretability at both the workflow level and model level.

At the workflow level, the agent explains every operational phase in detail, including debugging, pre-training, training, and inference. This trail allows clinicians to audit the model and pinpoint clinical reasoning errors. At the model level, the agent can explain DL algorithms or generate new tools that facilitate the interpretation of synthesized models. This includes autonomously integrating localized gradient-weighted class activation mapping (Grad-CAM) to localize image regions contributing to malignancy predictions and allowing direct visual correlation between model output and radiological features.

### 2.2 Controllability

The effectiveness of the radiological agent arises from the combination of its multi-disciplinary domain knowledge and the technical capabilities of its tools. Our approach establishes a flexible partnership where radiologists can control all the logic of the AI system with varying degrees of autonomy.

We tested the system across three levels of system autonomy (Figure 2). In the autonomous repair task, which represents the lowest level of human control and the highest level of agent autonomy, the agent can analyze an existing DL tool repository to identify missing components and regenerate the necessary components to restore functionality. The final output is then validated by a domain expert to eliminate hallucinations. In supervised improvement, an intermediate level of human control and agent autonomy, radiologists serve as supervisors to review and select improvement suggestions proposed by the LLM. This isolates the LLM’s capacity to pinpoint clinical reasoning errors and improve the DL tools relying only on its domain knowledge. For clinical customization, which involves the highest level of human control, radiologists specify desired clinical functionalities, and the LLM customizes the DL model to create new tools accordingly.

**Figure 2:**
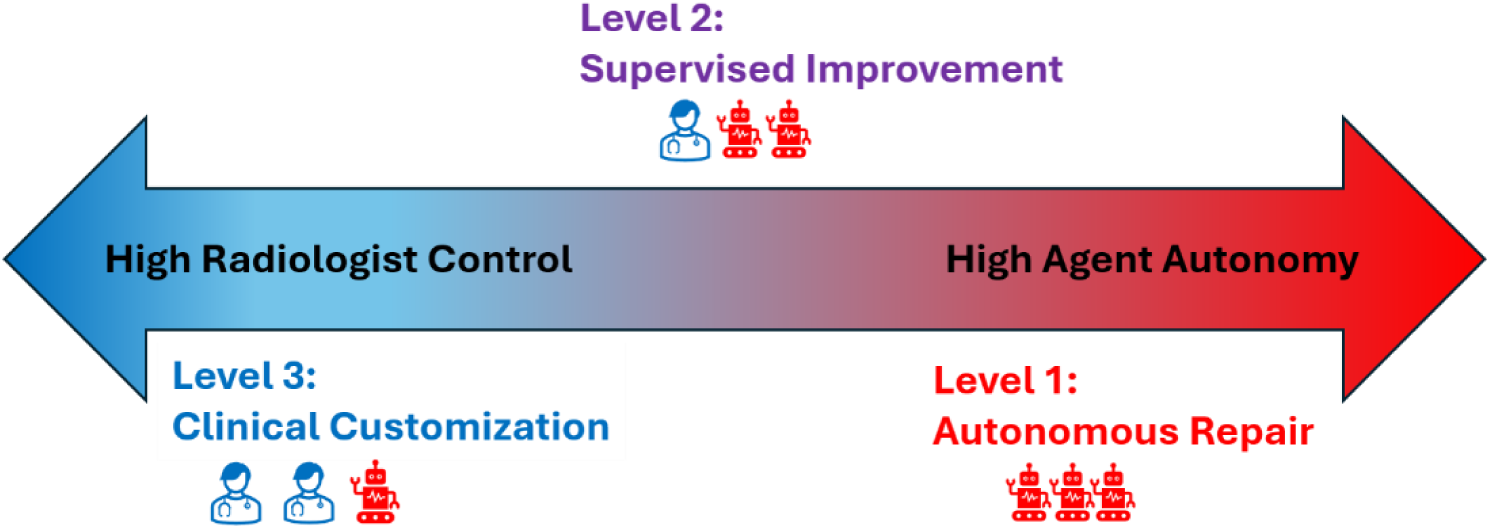
Three levels of radiologist controllability. Level 1 – Autonomous Repair: Lowest radiologist control and highest agent autonomy, where the agent can independently assess and repair the DL tools. Level 2 – Supervised Improvement: Moderate radiologist control and agent autonomy; the radiologist reviews and selects the improvements made by the agent. Level 3 – Clinical Customization: Highest radiologist control and lowest agent autonomy, where the radiologist directs the customization of DL tools.

### 2.3 Accessibility and Hardware Configuration

To facilitate the adoption of advanced radiological AI in resource-constrained settings, the AI workflow operates entirely within an open-source ecosystem, where all tasks can be run locally on a single consumer-grade computer (central PC) to minimize costs and cloud-related risks (more details are shown in Figure 1A). To protect data privacy, only de-identified queries may be utilized for external reasoning when necessary, making cloud-based LLMs optional.

We further implemented a distributed architecture, in which computationally intensive tasks, such as deep reasoning and model training, are assigned to a single consumer-grade central PC with an NVIDIA RTX 5090 GPU (approximate cost: $3,500–$5,000). This central PC delivers optimized model weights to multiple clinical interfaces that consist of standard PCs utilizing a lower-cost NVIDIA RTX 4070 GPU (approximate cost: $1,500–$2,000) for low-latency inference tasks.

## 3. Initialization Pipeline and Original Model Selection

The initialization pipeline began with the AI agent (OpenClaw) utilizing a search engine to identify candidate open-source DL repositories to serve as original models. The reasoning LLM (DeepSeek-R1) analyzed these candidates and generated natural language summaries outlining their workflow-level process and model-level interpretability for radiologists. From this search, we selected the 5th-place solution ^38^ from the 2023 Radiological Society of North America (RSNA) Breast Cancer Detection AI Challenge (as detailed in Appendix), hosted by the world’s largest radiology society, as our original model. According to the competition guidelines, predictions were generated at the breast level without localizing the lesion.

The original model was selected on the basis of the following considerations: the pre-training model in the 5th-place solution is missing in the open-source code repository, which enables us to evaluate the LLM’s capability for autonomous repairing. Furthermore, the original model ranked fifth among all submissions in the RSNA competition and therefore provides a well-established baseline to benchmark the LLM-optimized model against the public results from leading multi-disciplinary teams on the hidden RSNA test dataset.

## 4. Autonomous Repair

The agent mapped the target architecture by parsing the original codebase modules, configuration files, and documentation dependencies. It then executed an iterative agentic coding loop to test the candidate implementations of the missing pre-training model. After the LLM reconstructed the pre-training model by analyzing runtime error logs and adjusting syntax based on execution feedback, the resulting LLM-repaired system converged to generate DL weights (checkpoints) that reproduced the original 5th-place RSNA metrics.

## 5. Supervised Improvement

During the inference phase, the LLM identified a clinical reasoning flaw in the original DL model’s view aggregation mechanism. The original architecture utilized a standard arithmetic mean to aggregate scores across craniocaudal (CC) and mediolateral oblique (MLO) views. The LLM reasoned that this approach introduces a high risk of false negatives when a suspicious lesion is prominent in one view but obscured in another. To address this, the LLM proposed an additional “Max” pooling strategy in the DL model logic to ensure that strong positive single-view signals remain undiluted.

Our radiologists reviewed and confirmed the clinical validity of this architectural modification (Extended Data Figure 2 in Appendix). To isolate the possible impact from model training, we applied the original checkpoints to the LLM-optimized DL model. When evaluated against the RSNA hidden test dataset (5,415 patients), the model’s pF1 score increased from 0.52 to 0.56, elevating the model’s ranking from fifth place to surpassing all 1,687 submitted AI algorithms.

Following the validation framework established by McKinney et al. ^28^, we extended the evaluation of this LLM-optimized DL tool with both a retrospective study and a reader study.

## 6. Retrospective Study

In the retrospective comparison, we evaluated the LLM-optimized system against a public dataset from Emory University comprising 13,011 patients. Using biopsy-confirmed outcomes as the ground truth, we compared the AI’s predictions to the original clinical decisions made by radiologists. Under the U.S. standard of single-reader interpretation, radiologist performance was modeled using the Breast Imaging Reporting and Data System (BI-RADS) score assigned during the initial screening as a proxy for recall decisions (see Methods section ‘Interpreting clinical reads’ in ^28^). As illustrated in Figure 3, the AI system outperformed the mean radiologist, achieving statistically significant absolute improvements in AUC of 7.27% (95%CI:5.2-9.4%; *P*<0.001). Improved sensitivity at matched specificity, and vice versa, reflects a reduction in both false negative and recall rates.

**Figure 3.**
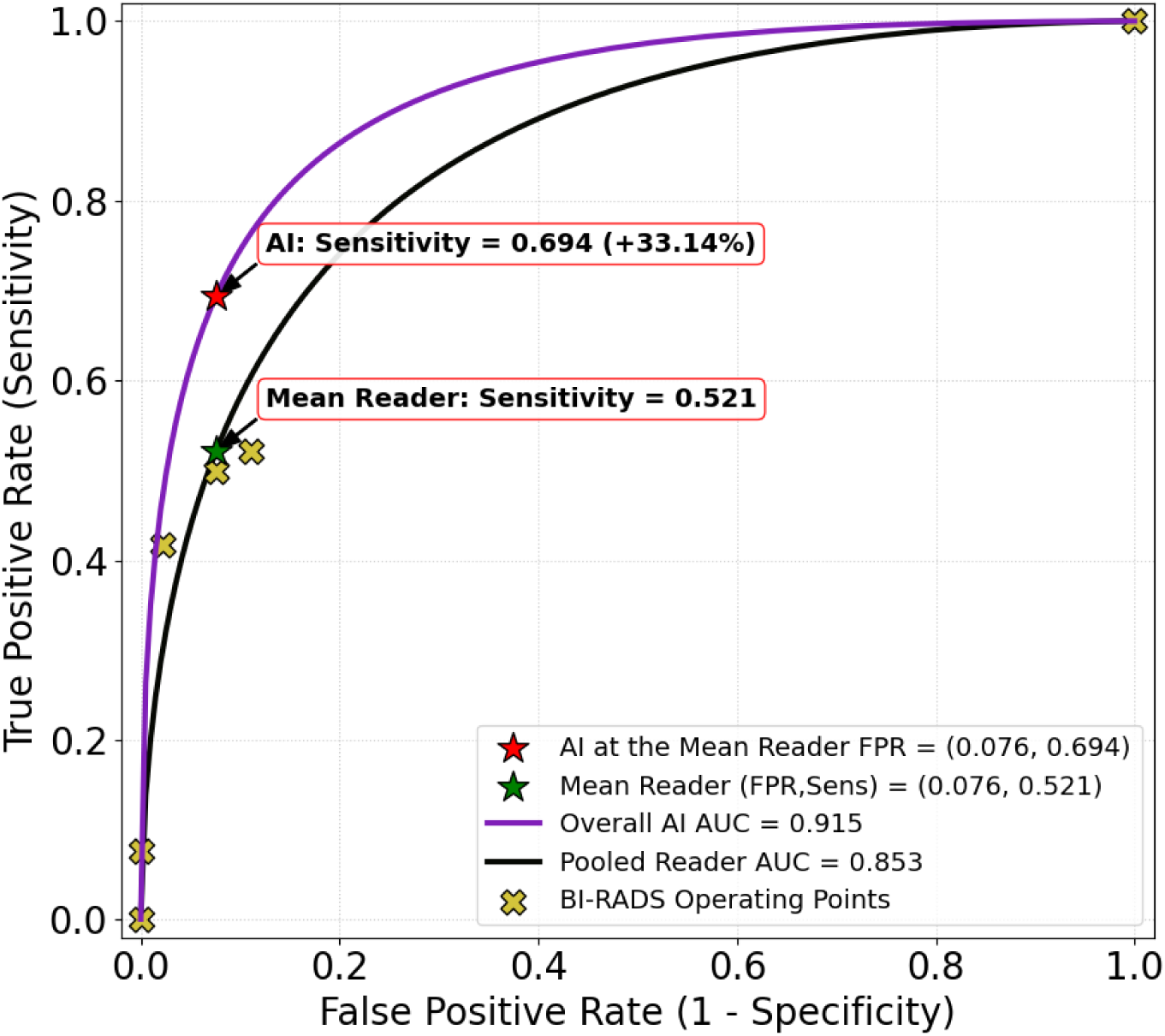
Performance of the AI system versus radiologists in breast cancer prediction. Receiver operating characteristic (ROC) curves illustrating the performance of the AI system (purple) and the mean radiologist (black) in the retrospective study with 13,011 patients. Cases were considered positive (n=344) if they received a biopsy-confirmed diagnosis of cancer within 2 years (27 months) of screening. The area under the curve (AUC) for the AI system (0.915) exceeds that of the pooled reader AUC (0.853), indicating improved overall discriminative performance. At the mean radiologist specificity of 0.924 (False Positive Rate: FPR = 0.076), the AI system (red star) achieves a sensitivity of 0.694, representing an absolute improvement of +33.1% (95%CI:24.2-41.5%; *P*<0.001) over the mean radiologist (green star; sensitivity = 0.521). Across the evaluated operating range, the AI system consistently demonstrates higher sensitivity for a given specificity, indicating a reduction in false-negative rates, while also achieving higher specificity at comparable sensitivity levels, potentially reducing screening recall rates.

## 7. Reader Study

We conducted a reader study involving six fellowship-trained, board-certified radiologists interpreting a total of 1,239 patients. The validation cohort consisted of three sub-populations (Dataset details are provided in Appendix):

**RSNA Sub-cohort** (500 patients): Sampled from the open training dataset for the RSNA mammography competition and interpreted by our radiologist team to calculate the human pF1 scores, which were then compared against our LLM-optimized DL model’s pF1 score generated from the RSNA competition pipeline with the hidden test set.

**US Emory Sub-cohort** (539 patients): Sampled from the Emory Breast Imaging Dataset (EMBED) to represent a challenging screening cohort with long-term longitudinal follow-up.

**China Sub-cohort** (200 patients): Sampled from the Chinese Mammography Database (CMMD) to evaluate cross-continental performance.

In the first phase of the reader study, four radiologists independently interpreted the RSNA sub-cohort, assigning binary cancer/no-cancer assessments at the breast level. We assumed the baseline patient characteristics of the RSNA training-pool sample to be epidemiologically representative of the hidden RSNA testing dataset. At a similar cancer prevalence of 4%, the pooled human readers achieved a pF1 score of 0.28 (95% CI: 0.26–0.30). The LLM-optimized DL model (pF1 = 0.56) outperformed the human reader team when evaluated under an identical RSNA competition pipeline (see Appendix section ‘RSNA Competition’).

The second phase of the reader study evaluated model generalizability across international sub-cohorts from US Emory and China sites. Six radiologists assigned a forced six-point BI-RADS assessment (BI-RADS 1/2, 3, 4A, 4B, 4C, and 5) for each breast, in order of increasing suspicion for malignancy. Following the methods described by McKinney et al. ^28^, performance metrics were analyzed across two clinical follow-up windows: a 1-year window (mean: 15 months) and a 2-year window (mean: 27 months), mapping to annual and biennial screening intervals. We included a 3-month operational buffer to account for real-world appointment scheduling variability and delays in the ascertainment of follow-up outcomes.

Cancer-positive ground truth was defined by a biopsy-confirmed histological diagnosis of malignancy within the specified follow-up window. True-negative labels required a minimum of one documented negative screening examination within that same longitudinal follow-up period. Cases lacking verified longitudinal tracking were excluded from the US Emory sub-cohort analysis. “Difficult cases” were defined as cases where the initial screening mammogram received a benign or negative interpretation and the breast cancer was diagnosed more than 1 year after that initial screening examination. The China sub-cohort was used to evaluate performance in a cohort without the “difficult cases”.

As detailed in Table 1, each participating radiologist evaluated a minimum of 100 patients. To capture inter-observer variability among radiologists with matched clinical experience levels, Reader 5 and Reader 6 interpreted the identical subset of cases. Additionally, Reader 1 and Reader 2 evaluated both the US Emory and the China datasets to assess performance variations across distinct patient populations and screening environments.

**Table 1.** Reader Study for the US and China datasets. Performance of the AI system was compared with that of radiologists of varying clinical experience across two independent datasets. In the US Emory cohort (follow-up window: 27 months), the AI achieved an AUC of 0.908, outperforming all readers, with the largest performance gap observed among those with fewer years of clinical experience. In contrast, radiologist performance in the China dataset (no longitudinal follow-up) was more comparable to AI (AUC = 0.917), with no statistically significant differences observed.

| **US Emory Data (follow-up windows of 27 months)** |  |  |  |  |
| --- | --- | --- | --- | --- |
| **Radiologist** | **Years in Practice** | **Difficult Case (%)** | **AUC** | **ΔAUC, (95%CI); *P* value**<br>ΔAUC = AUC(AI) - AUC(Reader) |
| Reader 1 | >10 | 12% - 13% | 0.647 | 0.326, (0.141, 0.450); *P*<0.001 |
| Reader 2 | >10 | 12% - 13% | 0.777 | 0.128, (0.042, 0.208); *P*=0.006 |
| Reader 3 | >10 | 7% | 0.892 | 0.015, (-0.025, 0.147); *P*=0.701 |
| Reader 4 | <5 | 1% | 0.613 | 0.248, (0.099, 0.445); *P*<0.001 |
| Reader 5 | <5 | 12% - 13% | 0.628 | 0.266, (0.090, 0.434); *P*=0.023 |
| Reader 6 | <5 | 12% - 13% | 0.508 | 0.386, (0.186, 0.467); *P*=0.018 |
| Pooled Reader AUC = 0.732; Overall AI AUC = 0.908;<br>ΔAUC = 0.176, 95%CI 0.112, 0.213; *P*<0.001 | | | | |
| **US Emory Data (follow-up windows of 15 months)** |  |  |  |  |
| Pooled Reader AUC = 0.761; Overall AI AUC = 0.916;<br>ΔAUC = 0.155, 95%CI 0.114, 0.209; *P*<0.001 | | | | |
| **China Data (no follow-up window)** |  |  |  |  |
| Reader 1 | >10 | 0 | 0.880 | -0.014, (-0.160, 0.053); *P*=0.746 |
| Reader 2 | >10 | 0 | 0.941 | -0.008, (-0.110, 0.059); *P*=0.798 |
| Pooled Reader AUC = 0.914; Overall AI AUC = 0.917;<br>ΔAUC = 0.003, 95%CI -0.108, 0.025; *P*=0.911 | | | | |

The LLM-optimized AI system exceeded the average performance of radiologists by a statistically significant margin, yielding an absolute increase in AUC of 0.176 (95% CI 0.112, 0.212; *P* < 0.001). As illustrated in Figure 4B and C, the AI system achieved higher sensitivity while maintaining equivalent specificity compared to pooled readers at both 15-month and 27-month follow-up intervals.

**Figure 4.**
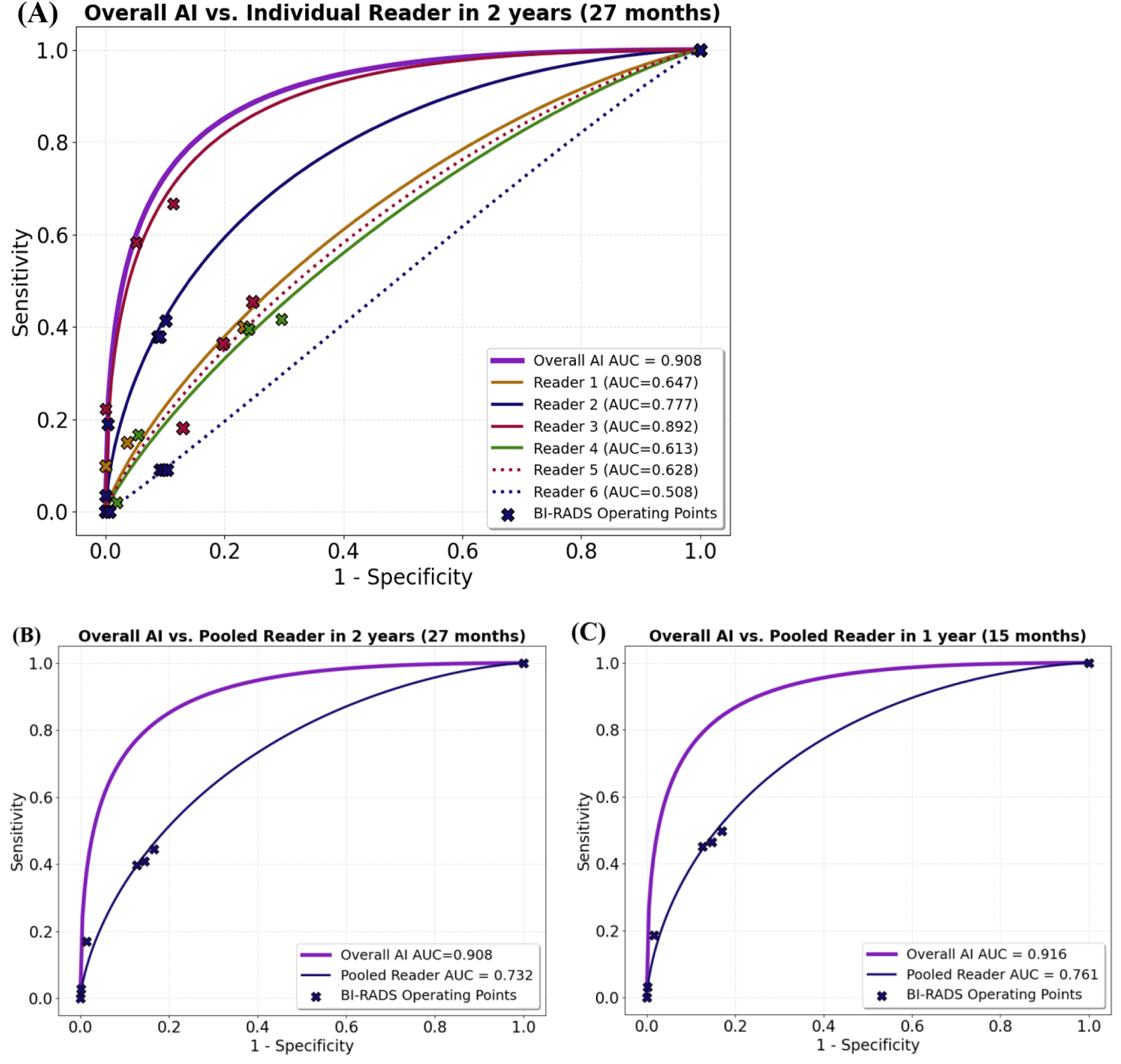
Performance of the AI model compared to independent readers. (A) Six readers rated each case/breast using a six-point BI-RADS scale (BI-RADS 1/2, 3, 4A, 4B, 4C, or 5). A fitted ROC curve for each of the readers is compared to the ROC curve of the AI model. Cases were considered positive (n=152) if they received a pathology-confirmed diagnosis of cancer within 27 months of the time of screening. (B) Pooled results (n=499) from all six readers using a 27-month interval for cancer definition. (C) Pooled results (n=438) from all six readers using a 15-month interval for cancer definition. Cases were considered positive (n=139) if there is a biopsy-proven malignancy within 15 months.

Stratified analysis showed no significant performance difference between AI and experienced radiologists when difficult cases were low in prevalence (*P* > 0.10), such as Reader 3 within the US cohort or Readers 1 and 2 within the China dataset. However, when difficult cases exceeded 12%, the AI system significantly reduced both false positives and false negatives compared with radiologists across all experience levels (*P* < 0.05).

## 8. Clinical Customization

The original DL model was limited to breast-level binary classification. In our study, we asked the agent to additionally output a continuous cancer probability with an adjustable decision threshold, while preserving the original binary classification. This additional score provides confidence information and allows the model to be tailored to different radiologists’ preferences, such as prioritizing sensitivity or specificity.

To further assist radiologist interpretation, the agent customized the original model through agentic coding (Figure 1.C) for localized heatmaps. Figure 5 illustrates this clinical utility in a representative discrepant case: a left breast cancer not detected by three out of four radiologists on the initial screening mammogram but diagnosed on the subsequent screening examination performed one year later. The AI-customized model correctly flagged the malignancy on the earlier examination, with the corresponding heatmaps localizing the subtle findings.

**Figure 5.**
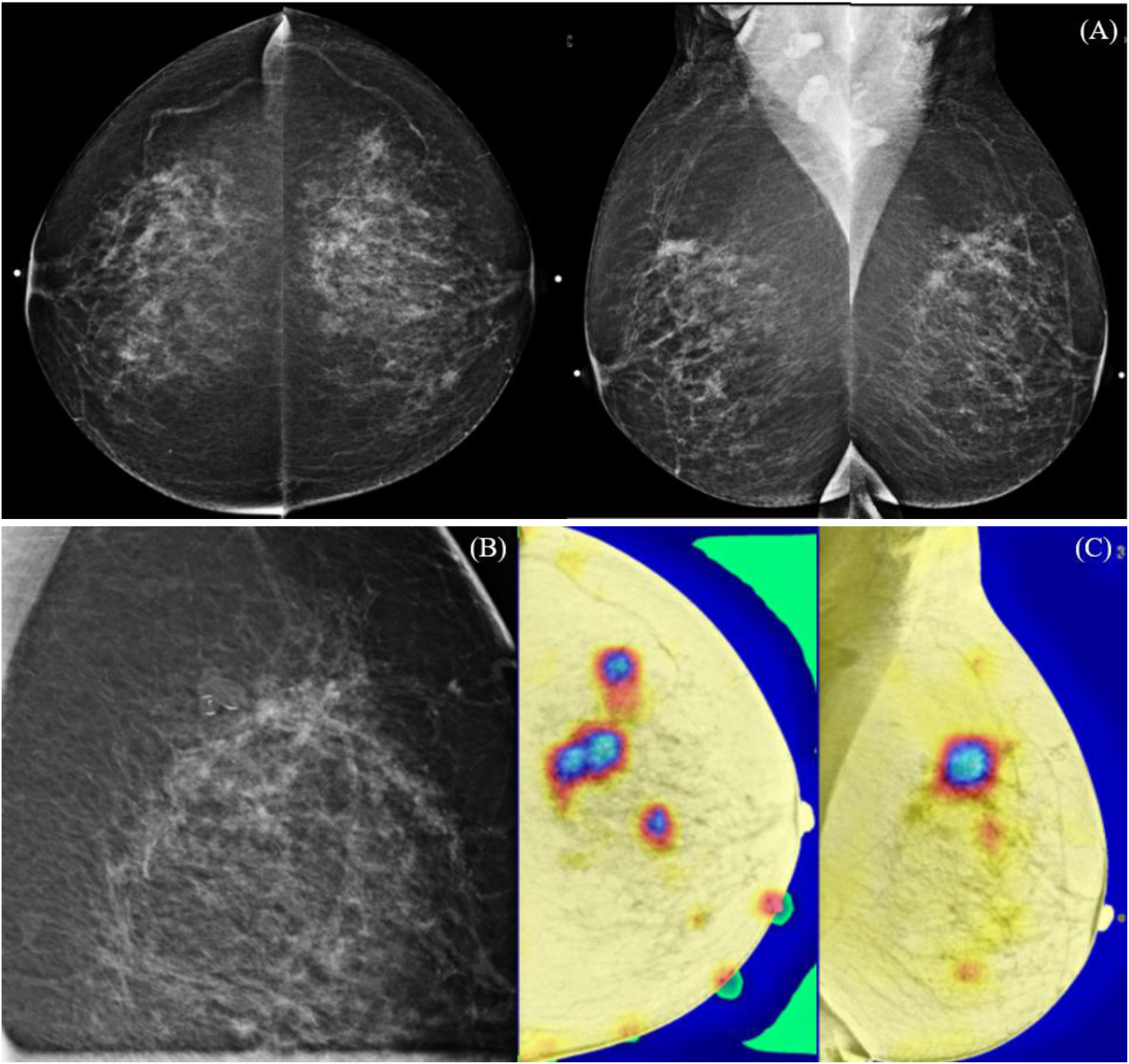
Discrepancies between radiologists and the AI model. (A) Screening mammogram views demonstrating a left breast cancer that was missed by three out of four radiologists but correctly identified by the AI model. (B) Magnification view showing the region of malignancy. (C) AI-generated heatmap highlighting regions of increased model attention, corresponding to the regions of malignancy.

We also tested the agent’s ability to retrain the DL tool with external datasets containing various data formats, allowing the model to be iteratively refined using institutional data that reflects local patient demographics.

## 9. Discussion

Recent advances in open-source AI, combined with the increasing capability of consumer-grade computers are transforming how radiological AI systems can be developed and deployed. Unlike the conventional approach, we introduce an LLM-driven agentic workflow built entirely on open-source components and executed on a single consumer-grade computer.

Using an open-source DL model from the RSNA Screening Mammography Breast Cancer Detection Challenge as a test case, we evaluated the workflow across increasing levels of agentic autonomy. At the highest level of autonomy (autonomous repair), the agent independently identified and reconstructed a missing pre-trained model in the public repository to reproduce the same pF1 score as the original model.

At the second level of agentic autonomy, the agent leveraged medical reasoning capabilities to identify a clinical reasoning flaw in the original model and introduced a multi-view consensus process that closely mirrors a human radiologist’s approach. The LLM-optimized model’s ranking is elevated from fifth place to outperforming all 1,687 submitted AI algorithms. This finding underscores that major performance gains can be achieved by optimizing clinical logic rather than developing entirely novel neural network architectures.

We validated the LLM-optimized model in a retrospective study using US dataset (n > 13,000 patients) alongside an independent reader study with international datasets (n>1,200). These complementary evaluations demonstrated the model’s potential clinical utility in reducing interval cancer and recall rates. While the LLM-improved model performs comparably to experienced radiologists in low-complexity cases, its performance advantage scales with case difficulty. In particular, the AI model identified malignancies on prior screening examinations that were not detected by radiologists. In the extended (27-month) follow up, the AUC for both AI and radiologists was lower than that at 15-month, as the extended interval makes cancer predictions more challenging. Nevertheless, the model maintained high performance, with an AUC exceeding 0.9 on both follow-up intervals.

Another implication of our findings is that the AI model may be capturing imaging features associated with future breast cancer development, rather than simply detecting cancers that were already evident at the time of screening. Despite extending the follow-up window to 27 months in our study, the LLM-optimized model maintained strong AUC. As a subset of these cancers were mammographically occult at the initial screening exam, the observed performance of the AI system under this extended follow up suggests potential value in future cancer risk prediction. If confirmed in prospective studies, such capability could support more personalized screening strategies.

At the third level of autonomy, the workflow demonstrated the capacity for clinician-directed customization. In response to radiologist feedback, the agent adapted existing tools for lesion localization and reconfigured training pipelines to accommodate diverse data structures from local clinical populations.

There are several limitations of this study. First, our evaluation relied on retrospective public datasets; prospective studies are required to quantify clinical performance within routine screening workflows. Second, the reader study was enriched for cancer cases and challenging cases and involved a limited number of readers. Broader validation across larger reader cohorts with varied image resolutions and digital breast tomosynthesis is an essential next step. Beyond augmenting radiologist performance, additional clinical utility of the AI system, such as in triaging cases, remains to be systematically evaluated.

In summary, our study demonstrates that an open-source, locally executable agentic workflow can deliver competitive radiological performance on consumer-grade hardware. This work serves as a proof-of-concept for a new paradigm in radiological AI, built upon explainability, controllability, and accessibility (Figure 1). By shifting toward low-cost, fully open-source, clinician-steered AI, this framework aims to democratize advanced AI in radiology and improve patient outcomes globally.

## Data Availability

All data produced in the present study are available upon reasonable request to the authors

https://www.rsna.org/artificial-intelligence/ai-image-challenge/screening-mammography-breast-cancer-detection-ai-challenge

https://www.kaggle.com/competitions/rsna-breast-cancer-detection/data

https://registry.opendata.aws/emory-breast-imaging-dataset-embed/

https://www.cancerimagingarchive.net/collection/cmmd/

https://physionet.org/content/vindr-mammo/1.0.0/

## Appendix (Methods)

### The Radiological Society of North America (RSNA) Competition

RSNA, the world’s largest radiology organization, hosted the Screening Mammography Breast Cancer Detection AI Challenge^39^ in 2023. More than 1,600 teams took part, making it one of the most widely participated RSNA AI challenges to date. The competition aimed to advance AI tools that support cancer detection in screening mammography.

In total, 1,687 algorithms were submitted from teams around the world. Because many of these models were released as open source, the Challenge created a large, shared resource for the community. The availability of both open-source algorithms and a carefully curated imaging dataset enable benchmarking and fosters continued innovation in the development of mammography AI systems for both research and clinical applications. All submitted models generated breast-level predictions.

### RSNA Dataset

Screening mammograms were collected from two clinical sites: Emory Healthcare (Atlanta, Georgia, USA), and BreastScreen Victoria (Victoria, Australia). All examinations were de-identified according to HIPAA standards and approved by the respective institutional review boards. Eligible cases were routine screening mammograms from asymptomatic women. Only studies with all four standard screening views (bilateral CC and MLO) and adequate image quality were included. No age restrictions were applied; the age distribution reflected the screening practices at each site.

Screening typically begins at age 40 at Site 1, though younger high-risk patients may also be screened and women with a prior history of breast cancer are not excluded. At Site 2, screening generally begins at age 50, but women aged 40–49 may participate after discussing screening with their primary care physician.

Cancer and benign lesions were confirmed pathologically. Benign/negative cases that did not undergo biopsy required at least one year of negative follow-up, and no interval cancers were included. To ensure enough positive cases for model development, the dataset was enriched to a 4% cancer prevalence. From Site 1, 10,000 participants (including 400 cancers) were sampled from the Emory Breast Imaging Dataset (EMBED, 2013–2020). From Site 2, 10,000 participants with 400 cancers from 2013–2015 were selected. Only one set of mammograms were available per patient, so long-term outcome (e.g., 27-month intervals) could not be established in this dataset.

The hidden evaluation set contained 5,415 patients. The median screening age was 59 years (IQR 52–66). Images were acquired primarily on Hologic systems (58.2%), followed by Siemens Healthineers (17.2%), Philips (16.5%), GE HealthCare (6.3%), and Fujifilm (1.8%). Algorithms were ranked using a probabilistic F1 metric on this hidden test set; the top model achieved a score of 0.55.

Competition participants were also given access to a separate training dataset of 11,913 patients (including 486 cancer patients) drawn from the same two sites. This training set did not overlap with the hidden evaluation set. Teams were free to supplement the provided data with additional public datasets such as VinDr-Mammo and the Digital Database for Screening Mammography (DDSM), described in later sections.

### US Emory Dataset

The Emory BrEast Imaging Dataset (EMBED) ^40^ comprises mammography examinations acquired across four hospitals in the Emory Healthcare system over an eight-year span (2013–2020). Image acquisition was predominantly performed on Hologic systems (92%), with GE Healthcare (6%) and Fujifilm (2%) accounting for the remainder. The patient population is racially and ethnically diverse, including approximately 40% White, 40% African American, 6.5% Asian, and 5.6% Hispanic.

In total, EMBED includes approximately 364,000 mammogram examinations from roughly 110,000 patients, along with about 60,000 annotated lesions tied to structured descriptors and biopsy-confirmed pathology. All examinations included in this study were standard 2D digital screening mammograms consisting of CC and MLO views of both breasts and were interpreted by board-certified, fellowship-trained breast imaging radiologists.

A small portion of EMBED (approximately 5,900 patients) was used in the RSNA training dataset. The US Emory Dataset analyzed in our study is a public open-data subset representing about 20% of the full cohort. It includes 20,393 patients in screening mammogram examinations. Among those, 453 patients had biopsy-confirmed breast cancer. This corresponds to a cancer prevalence of 2.22% (breast-level cancer prevalence: 1.17%), which was used for inverse probability weighting in the statistical analysis for both the retrospective study and the reader study.

Cancer status was assigned using biopsy results or longitudinal follow-up. If either breast had a biopsy-proven malignancy, the patient was considered cancer-positive. Because many patients returned for multiple screening rounds, the dataset naturally forms a multiyear series rather than a single-timepoint snapshot. Patients with mammograms considered benign or negative needed at least one subsequent negative screening exam within either the 15-month or 27-month follow-up windows.

To establish the evaluation cohorts, we excluded patients with insufficient follow-up inadequate image quality, or overlap with the RSNA training set. After these exclusions, the 27-month cohort comprised 13,011 patients, including 344 cancer patients. The 15-month cohort comprised 11,591 patients, including 339 cancer patients.

### China Dataset

The China Dataset comes from the publicly available China Mammography Database (CMMD) ^41^. It includes 1,775 patients who underwent mammography between July 2012 and January 2016 (mean age 47.6 years; range 18–87). Imaging was performed at two clinical sites: Sun Yat-sen University Cancer Center in Guangzhou and the Nanhai Affiliated Hospital of Southern Medical University in Foshan. Exams were acquired on GE Senographe DS and Siemens Mammomat Inspiration systems, following a consistent acquisition protocol. Each patient had both standard CC and MLO views.

Two of the radiologist readers, each with more than ten years of experience, interpreted the mammograms using the American College of Radiology’s BI-RADS framework. Of the 1,775 patients, 1,310 (73.8%) had biopsy-proven breast cancer and 465 (26.2%) had benign pathology. Cancer status was assigned at the patient-level—if either breast showed malignancy, the patient was considered positive. The age distribution spanned early adulthood through older age, with most patients falling between 40 and 59 years.

This dataset was used solely as an external test set to evaluate model performance and assess generalizability across continents. No images from CMMD were used for model training or hyperparameter tuning. Because only a single exam per patient is available, long-term follow-up windows—such as the 12-month and 27-month intervals used in the US Emory dataset—cannot be assessed here.

### Pre-training Dataset

For the pre-training stage, we adopted an approach similar to the fifth-place “Racers” team in the RSNA competition and made use of the public VinDr-Mammo dataset ^42^. This dataset contains 20,000 digital mammograms from 5,000 patients, collected on full-field digital systems at two major hospitals in Hanoi, Vietnam. Each exam includes the standard bilateral CC and MLO views. A notable strength of VinDr-Mammo is the level of expert annotation: every study was double-read by radiologists, who assigned breast-level BI-RADS categories, density ratings, and lesion classifications.

In our workflow, VinDr-Mammo served only as a pre-training resource, not fine-tuning or evaluation. This follows the strategy demonstrated by the Racers team in the 2023 RSNA Screening Mammography Challenge, who showed that models benefit from being initialized on VinDr-Mammo’s rich annotations before being trained on the main RSNA training dataset.

According to the instructions from the 5^th^ solution about pre-training, we converted VinDr’s lesion bounding boxes into image-level multi-label lesion annotations, discarding the coordinates. Both backbone networks were then trained in a multi-task setup that included BI-RADS prediction, breast density classification, and lesion-type classification. This stage served purely as model initialization before entering the main training pipeline.

One practical complication is worth noting: the pre-training model from the fifth-place solution was missing from its public repository, and had to be reconstructed from the available code and documentation. Our AI scientist confirmed this omission and used it to test our proposed AI system’s autonomous repair feature.

### Additional Training Dataset

Following the approach used by the fifth-place team in the RSNA competition, we incorporated Mini-DDSM into our primary training pipeline as an additional public dataset. The Mini-DDSM dataset is a cleaned and modernized version of the original Digital Database for Screening Mammography (DDSM), which was in outdated compression formats and had to be reprocessed. The studies were contributed by four U.S. clinical centers—Massachusetts General Hospital, Wake Forest University School of Medicine, Sacred Heart Hospital, and Washington University School of Medicine.

These films were later digitized using four scanners common at the time: DBA, Howtek, Lumisys, and VIDAR. Disease status was determined through radiologic review and confirmed by biopsy for suspicious findings. The dataset includes 679 cancer patients and 1,273 non-cancer patients, for a total of 1,952 patients. This corresponds to a cancer prevalence of 34.8%, which is far higher than in routine screening populations.

### The Fifth Place Solution in the 2023 RSNA Competition

#### Stage 1: Multi-Task Pre-training

The training pipeline starts with a multi-task pre-training stage built on the VinDR-Mammo dataset, which provides a base for feature extraction. For this step, the original bounding-box annotations are simplified into image-level labels, and the coordinate information is dropped. The standard timm backbones (EfficientNet: v2s and b5) are used only as feature extractors—their classification layers are removed so that the network outputs clean global features. These features feed into three newly initialized linear heads trained in parallel. Each head targets a different clinical label set: a five-class BI-RADS category (levels 1–5), a four-class breast density category (A–D), and an eleven-label vector covering common lesion types such as masses and suspicious calcifications (Extended Data Figure 1).

**Extended Data Figure 1.**
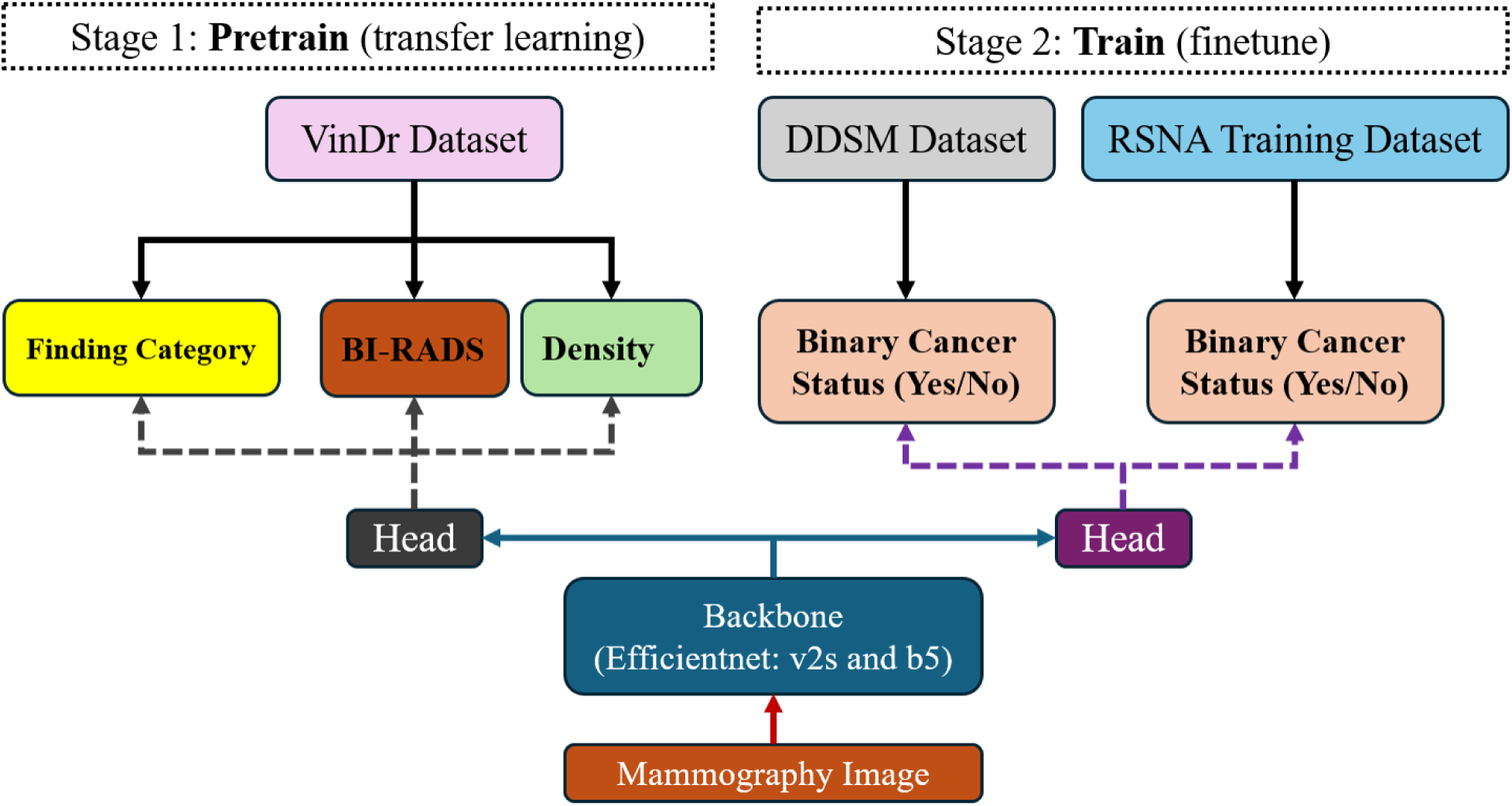
Two-stage training process. Stage 1 (Pretraining) uses multiple feature types (Findings category, BI-RADS classification, and Density) from the VinDr dataset. Stage 2 (Training) uses those weights from the pretrain to perform binary cancer classification with the Mini DDSM and RSNA training datasets.

#### Stage 2: Fine-Tuning (Training)

Once pre-training is complete, the final checkpoints are loaded and fine-tuning begins on a combined dataset made up of the RSNA training dataset and the DDSM dataset. Because breast cancer screening data is extremely imbalanced, the model is reconfigured into a deep-supervision setup, adding auxiliary loss heads to earlier EfficientNet blocks to stabilize learning. Balanced batch sampling is enforced throughout training to keep the network from collapsing into all-negative predictions. The image pipeline applies a mix of augmentations—shift/scale/rotate, horizontal and vertical flips, brightness and contrast changes, and coarse dropout. For the main optimization objective, standard Binary Cross-Entropy (BCE) loss was utilized.

#### Stage 3: Inference and Hierarchical Decision Aggregation

At inference time, predictions are produced through a two-stage hierarchical process that blends model-level ensemble with breast-level clinical reasoning. The first stage operates at the image level. Each mammogram is run independently through all ten fine-tuned models—the five cross-validation folds of both EfficientNet_v2s and EfficientNet_b5. Their outputs are averaged to produce a single malignancy probability for each image.

The second stage shifts from individual images to breast-level interpretation. A single breast typically has multiple views (most often CC and MLO), so each breast side accumulates several image-level scores. These predictions are grouped by patient and laterality, and the final malignancy probability for each breast is taken as the mean across all available views (Mean score comparison). In our study, the LLM generated improvement (Max score comparison) was added in this stage after approval from the radiologists (Extended Data Figure 2).

**Extended Data Figure 2.**
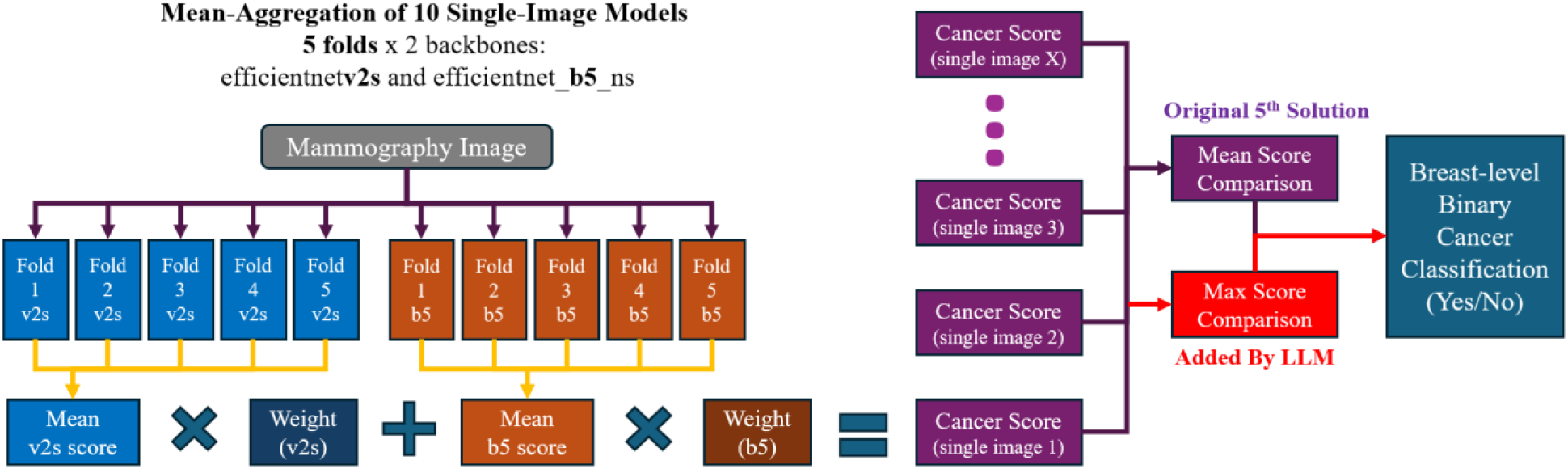
Stage 3 (Inference). Each mammogram is evaluated by an ensemble of ten fine-tuned models (five EfficientNet-v2s and five EfficientNet-b5 folds). Image-level cancer probability is calculated from the ten fine-tuned models. The final breast-level binary cancer classification (cancer versus non-cancer) is calculated from the comparison between the mean (or max) score and the user-defined thresholds.

### Statistical analysis

In the first phase of the reader study using the RSNA sub-cohort, we enriched it to a breast-level cancer prevalence of 30.1% to maximize statistical power for detecting subtle performance differences without requiring an impractically large reader caseload. We then applied inverse probability weighting (IPW), upweighting the normal cases to mirror the original cancer prevalence of the full RSNA open training dataset of 4.08% (breast-level of 2.06%) ^28^. We assumed the baseline patient characteristics of the RSNA training-pool sample to be epidemiologically representative of the hidden RSNA testing dataset.

The second phase of the reader study evaluated model generalizability across international sub-cohorts from US Emory and China sites. Similar to the first phase, cancer prevalences were enriched for both the US Emory and China sub-cohorts (breast level of 16.38% and 30.97%, respectively). To obtain unbiased performance estimates for both radiologists and AI within a more realistic screening population, we then applied IPW across all statistical analyses (including the retrospective study) using the original cancer prevalence (2.22%) of the raw US Emory dataset as a reference (The raw China dataset cancer prevalence of 73.8% is too high and thus unsuited as a reference).

We calculated confidence intervals for the performance differences using bootstrapping with 1,000 replications and evaluated statistical significance using a permutation test. For the permutation test, we ran 10,000 trials where the reader and AI scores were randomly swapped for each case to construct a null distribution of radiologist-AI differences. We then determined a two-sided *P*-value by comparing our observed performance statistic against the empirical quantiles of this randomized distribution.

During the reader study using the Emory and China datasets, radiologists graded cases under a forced BI-RADS classification (similar to ^28^) where BI-RADS 0 was not allowed. This yielded a 6-point index of suspicion for malignancy. We collapsed scores of BI-RADS 1 and 2 into a single baseline category of lowest suspicion, while treating scores of 3, 4A, 4B, 4C, and 5 as separate, increasing levels of suspicion. Because the BI-RADS operating points did not reach high-sensitivity levels, we avoided potential biases from non-parametric analysis by fitting parametric ROC curves to the data using a binormal model. We prioritized these BI-RADS-based scores due to their clinical relevance in real-world screening. Similarly, we fitted a parametric ROC curve to discretize AI system scores. We then compared the AI system’s performance against the radiologist panel in reader studies. All statistical computations, including *P*-values and confidence intervals, were implemented in Python using the NumPy and SciPy packages.

## Author contributions

L.C. and T.H. contributed to the conception, the study design, and the first draft of the manuscript; J.H.P., J.C.H., T.H., K.M., U.I., S.S., V.T., N.Y., F.K.A., A.R.M., J.G.S. and B.E.D. contributed to interpretation of the data; L.C., T.H. and S.H. contributed to data analysis; all authors reviewed and contributed to the final manuscript.

